# Inpatient outcomes for hospitalized older adults with rhinovirus

**DOI:** 10.1101/2021.04.09.21255232

**Authors:** Olivier Del Corpo, Emily G. McDonald, Luisa Smyth, Elizabeth Smyth, Moneeza Walji, Matthew P. Cheng, Charles Frenette, Ramy R. Saleh, Todd C. Lee

**Affiliations:** Faculty of Medicine, McGill University, Montreal, Canada; Clinical Practice Assessment Unit, Department of Medicine, McGill University, Montréal, Canada; Division of General Internal Medicine, Department of Medicine, McGill University, Montréal, Canada; Division of Hematology, Department of Medicine, McGill University, Montréal, Canada; Division of Infectious Diseases, Department of Medicine, McGill University, Montréal, Canada; Division of Medical Oncology, Department of Medicine, McGill University, Montréal, Canada

**Author notes:** **Corresponding author:** Olivier Del Corpo, McGill University, 845 Sherbrooke St W, Montréal, QC Canada H3A 0G4.

**Keywords:** Rhinovirus, mortality, hospitalization, prognosis, outcomes

## Abstract

**Background:** Rhinoviruses account for many cases of the “common cold” and infection is often self-limiting. As such, there is a lack of data regarding the inpatient outcomes of individuals hospitalized with rhinovirus infection. Given the generalized poorer prognosis of elderly admitted with respiratory viral infections, we assessed the mortality rate of general medical patients admitted with rhinovirus infection along with the major risk factors associated to mortality.

**Methods:** We performed a retrospective chart review of patients admitted to our clinical teaching ward from December 2013 to June 2017.

**Results:** Overall, 12.5% of patients admitted with rhinovirus infection died within 90 days of admission. The median age of admitted patients was 70 years-old. In univariable analysis, age (OR 1.05; 95% confidence interval (CI) 1.01-1.09) and the need for oxygen at presentation (OR 3.23; 95% CI 1.06-9.86) were associated with death while obstructive pulmonary disease or asthma (OR 0.10; 95% CI 0.01-0.81) was associated with survival. In the multivariable model, age (aOR 1.04; 95% CI 1.00-1.09) and obstructive lung disease (aOR 0.09 95%CI 0.01-0.73) remained significant whereas the requirement for oxygen at presentation did not (aOR 2.78; 95% CI 0.84-9.23).

**Conclusion:** Our study reveals that rhinovirus is an important cause of both morbidity and mortality in the elderly and further highlights the need for studies of potentially effective treatment options. In the meantime, we suggest that rigorous respiratory hygiene measures and quality older adult care should be practiced when caring for at-risk adults.

## BACKGROUND

Rhinoviruses are best known for causing a significant proportion of non-influenza viral respiratory tract infections. With the exception of multimorbid or immunocompromised hosts, infections are often self-limiting and no antiviral treatment is available. Infections with rhinoviruses rarely result in severe disease or lead to hospitalization (1). As such, there is a lack of data regarding the outcomes of patients hospitalized with these viruses. We sought to look at hospitalized patients with confirmed rhinovirus infections and characterize the major risk factors associated with in hospital mortality. We also wanted to compare and contrast rhinovirus in medical inpatients with influenza and severe acute respiratory syndrome coronavirus 2 (SARS-CoV-2).

## METHODS

Patient data were collected from the McGill Clinical Teaching Unit Cohort, a retrospective cohort of all consecutive admissions to our 52-bed medical clinical teaching unit in Montreal, Canada from December 2013 to June 2017. Patients were considered rhinovirus positive if they had a nasopharyngeal swab in the emergency department or within the first 48 hours of admission which was positive by polymerase chain reaction (PCR) for rhinovirus (2). Only the first eligible admission per patient was considered. Patients were excluded only if there was a lack of information on their comorbid conditions.

Categorical variables were compared using the Chi-square test. Continuous variables were expressed as median and interquartile range (IQR) and compared with the Wilcoxon rank-sum test. Variables with p < 0.05 in the single variable analysis were considered for the multivariable logistic regression analysis. To avoid overfitting, we limited our model to 3 predictor variables. Statistical testing was conducted using STATA version 15.1.

## RESULTS

A total of 4,103 patients were admitted from December 2013 to July 2017. Of those, 126 patients (3.1%) tested positive for rhinovirus via RT-PCR at the time of presentation. Of the 120 with complete data (see Table 1), 64 (53.3%) patients were men. The median age of patients admitted with rhinovirus was 70 years old (IQR: 54.5-80). The most common comorbidity was hypertension (46.7%), followed by chronic obstructive lung disease (COPD)/asthma (36.7%). Despite the viral PCR result, most (76.7%) patients received antibiotics.

**TABLE 1.** Patient demographics and clinical characteristics^a^

|  | **Total**<br>**(n = 120)** | **Survived**<br>**(n = 105)** | **Deceased**<br>**(n = 15)** |
| --- | --- | --- | --- |
| **Demographics** |  |  |  |
| Age, median (IQR), years<sup>c</sup> | 70 (54.5-80) | 68 (53-78) | 79 (66-87) |
| Male, n (%) | 64 (53.3) | 54 (51.4) | 10 (66.7) |
| Female, n (%) | 56 (46.7) | 51 (48.6) | 5 (33.3) |
| **Admission:** |  |  |  |
| Pneumonia | 30 (24.8) | 35 (33) | 5 (33.3) |
| COPD/Asthma exacerbation | 23 (19) | 23 (21.7) | 0 (0) |
| Upper respiratory tract infection | 12 (9.9) | 11 (10.3) | 1 (6.6) |
| Other | 56 (46.2) | 47 (44.3) | 9 (60) |
| Length of stay, median (IQR), days<sup>c</sup> | 5.5 (3-13) | 5 (3-13) | 15 (4-20) |
| **Comorbidities** |  |  |  |
| Hypertension | 55 (46.7) | 47 (44.8) | 8 (53.3) |
| CHF | 14 (11.7) | 10 (9.5) | 4 (26.7) |
| Active cancer | 40 (34.2) | 33 (31.4) | 7 (46.7) |
| Diabetes type 2 | 32 (26.7) | 30 (28.6) | 2 (13.3) |
| COPD/Asthma<sup>c</sup> | 44 (36.7) | 43 (41) | 1 (6.7) |
| Atrial fibrillation | 24 (20) | 21 (20) | 3 (20) |
| CAD/Myocardial infarct | 25 (21.7) | 22 (21.9) | 3 (20) |
| Cirrhosis | 5 (4.2) | 3 (2.9) | 2 (13.3) |
| **Symptoms** |  |  |  |
| Cough (n = 113) | 90 (79.6) | 78 (78.8) | 12 (85.7) |
| Sputum (n = 112) | 58 (51.8) | 50 (50.5) | 8 (57.1) |
| Shortness of breath (n = 116) | 78 (69.6) | 67 (66.3) | 11 (73.3) |
| Chest pain (n = 110) | 22 (20) | 22 (22.7) | 0 (0) |
| Vomiting (n = 110) | 10 (9.1) | 9 (9.3) | 1 (7.7) |
| Diarrhea (n = 111) | 11 (9.9) | 9 (9.3) | 2 (14.3) |
| **Vitals** |  |  |  |
| Temperature > 38°C (n = 109) | 19 (17.4) | 16 (16.8) | 3 (21.4) |
| Oxygen saturation (n = 115),<br>median (IQR), % | 96 (93-98) | 96 (93-98) | 96 (94-97) |
| Oxygen required<sup>c</sup> (n = 116) | 42 (35.9) | 33 (32.7) | 9 (60) |
| **X-Ray** |  |  |  |
| Received (n = 114) | 114 (95) | 100 (95.2) | 14 (93.3) |
| No pneumonia | 81 (71.1) | 70 (70) | 11 (78.6) |
| Pneumonia | 33 (28.9) | 30 (30) | 3 (21.4) |
| **Antibiotics** |  |  |  |
| Received | 92 (76.7) | 80 (76.2) | 12 (80) |
| Not received | 28 (23.3) | 25 (23.8) | 3 (20) |

<sup>a</sup> All Data are given as number (percentage) unless otherwise indicated.
<sup>b</sup> $P < 0.01$ ; <sup>c</sup> $P < 0.05$ .
Abbreviations: IQR, interquartile range; CHF, congestive heart failure; COPD, chronic obstructive pulmonary disease; CAD, coronary artery disease

Overall, fifteen patients (12.5%) died within 90 days of admission, of whom 10 (66.6%) were men. The majority of deaths were early with 11 deaths (9%) within 30 days. 8 of 15 deaths were due to pneumonia or related respiratory failure; 1 was due to *C. difficile*; and the remainder were due to cardiac, oncologic, or other causes. In univariable analysis, age (OR 1.05; 95%CI 1.01-1.09) and the need for oxygen at presentation (OR 3.23; 95%CI 1.06-9.86) were associated with death while obstructive pulmonary disease or asthma (OR 0.10; 95% CI 0.01-0.81) was associated with survival. Otherwise, there were no significant differences. In the multivariable model, age (aOR 1.04; 95% CI 1.00-1.09) and obstructive lung disease (aOR 0.09 95%CI 0.01-0.73) remained associated with death (p < 0.05) whereas the requirement for oxygen at presentation was no longer statistically significant (aOR 2.78; 95% CI 0.84-9.23).

## DISCUSSION

Viral respiratory tract infections are among the leading causes of morbidity and mortality worldwide (3). While influenza and more recently SARS-CoV-2 have been well recognized as an important cause of hospitalization and subsequent mortality in older adults, the roles of other respiratory viruses such as rhinoviruses remain less well defined (4). Rhinoviruses account for over 30% to 50% of common colds yearly. There are over 100 serotypes of human rhinoviruses which can cause respiratory tract infections in every age group and recurrent infections are common (5). With the exception of severely immunocompromised patients, infection with rhinoviruses are thought to be relatively self-limiting and benign. However, recent studies have demonstrated high associated mortality, specifically in older adults (6). Unfortunately, given the number of serotypes and lack of therapeutics targeting rhinoviruses directly, current medical management remains supportive.

In this cohort study spanning 5 years, we found that 12.5% of rhinovirus-infected patients admitted to our medical teaching unit died within 90 days of presentation. The major risk factor associated with mortality was age. This is in concordance with other inpatient studies which have found a rhinovirus-associated mortality of 8.3% (4) and long-term care facility studies which have demonstrated a mortality rate ranging from 6 to 21% in elderly patients (6, 7). These results further exemplify the poorer outcomes in elderly inpatients with respiratory virus infections and magnifies the need for appropriate prevention, support, and treatment.

For the majority of respiratory viruses, including influenza virus and SARS-CoV-2, the presence of chronic underlying illnesses are risk factors for severe outcomes in adults (8, 9). Although there was no significant difference in the majority of comorbidities between alive and deceased patients, our patients are polymorbid and the number of events was low compared to the number of predictors. Our findings are consistent with other studies that suggest increased morbidity from rhinovirus infection in patients with chronic underlying illnesses (1).

When adjusting for age, the mortality rates of inpatients infected with rhinovirus are comparable to that of influenza virus and common coronaviruses. One of the largest retrospective nationwide studies analyzing six influenza seasons in Spain demonstrated a mortality rate of 8.2% in patients over the age of 65 admitted with influenza pneumonia (10). Furthermore, outbreaks in long-term health care facilities have demonstrated mortality rates as high as 8.4% in patients infected with coronavirus strains 229E and OC43, which are responsible for the common colds (5). By comparison, inpatient mortality for SARS-CoV-2 in older medical inpatients was approximately 16.5% in a recent pan-Canadian cohort (SPRINT-SARI, S. Murthy Personal Communication, July 24, 2020).

As our results highlight, patients admitted with rhinovirus have a notable risk of short-term mortality. Some of these deaths will be due to the comorbid illnesses which were the real cause for admission to hospital (e.g. advanced cancer, dementia), some will be due to decompensation of chronic medical conditions like congestive heart failure or renal disease, and some will be due to the functional decline and iatrogenic harm of hospitalization which occurs in older adults. However, it can be challenging to attribute the cause of death in these complex cases and some of the deaths may be due to rhinovirus itself. In addition to optimal supportive care and multidisciplinary approaches like the “ACE-unit” (11), it is interesting to consider the role of other treatment options. Currently, dexamethasone has demonstrated promising results in SARS-CoV-2 infected patients who require oxygen (9). While steroids carry the risk of side effects in older adults, it is tempting to consider their role in rhinoviral pneumonitis. No randomized controlled trials have studied the effects of oral or inhaled corticosteroids in hypoxic influenza or rhinovirus infected patients. Indeed, the negative association of patients with chronic obstructive pulmonary disease and mortality in this cohort may have been mediated through the use of steroids for COPD exacerbation. Corticosteroids like budesonide have demonstrated antiviral and anti-inflammatory activity in rhinovirus *in vitro* and in mice further suggesting that steroids may be a candidate for study in the treatment of human rhinovirus infection (12).

Our study does have several limitations. Our findings are from a single Canadian tertiary care hospital and may not be generalizable to other hospitals and countries. Furthermore, the population of patients admitted to a clinical teaching include those with multiple comorbidities which may result in an elevated mortality rate. Moreover, patients were tested for rhinovirus by RT-PCR; however, agents isolated from the nasopharynx can represent asymptomatic colonization rather than pathogenic infection. Also, as testing is a clinical decision, only those patients where the treating team specifically looked for respiratory viruses were included. Finally, as previously discussed, attribution of mortality is extremely challenging. However, 30 patients were admitted with a diagnosis of pneumonia, 23 with COPD or asthma exacerbation and 12 with upper respiratory tract infection so for a large number of these patients the rhinovirus seems to have been directly contributory. The strengths of our study are the large overall sample size of admitted patients with rhinovirus and the longitudinal nature over several years such that the results are not restricted to a single circulating strain.

## Conclusion

Although rhinovirus commonly causes mild symptoms in healthy individuals, it can be an important cause of both morbidity and mortality in the elderly with similar overall mortality to hospitalizations involving influenza and other “common cold” viruses. Larger studies are needed to fully characterize the nature of rhinovirus infection in the elderly and there is a need for a study of potentially effective treatment options. In the meantime, respiratory hygiene measures, as for SARS-CoV-2 and quality older adult care emphasizing avoidance of iatrogenesis should be practiced when caring for at-risk older adults given the ubiquitous nature of these respiratory viruses and the associated rate of mortality.

## Data Availability

Data is available upon request

## Acknowledgements

Not applicable.

